# Self-reported real-world safety and reactogenicity of COVID-19 vaccines: An international vaccine-recipient survey

**DOI:** 10.1101/2021.02.26.21252096

**Authors:** Alexander G. Mathioudakis, Murad Ghrew, Andrew Ustianowski, Shazaad Ahmad, Ray Borrow, Lida Pieretta Papavasileiou, Dimitrios Petrakis, Nawar Diar Bakerly

## Abstract

**Background:** The safety of COVID-19 vaccines has been demonstrated in selected populations in recent studies, but more data in specific groups is needed to inform vaccine choice and health policy.

**Objectives:** An international, online survey was conducted to compare the safety, tolerability and reactogenicity of available COVID-19 vaccines in different recipient groups.

**Methods:** This survey was launched in February 2021, for 11 days. Recipients of a first COVID-19 vaccine dose ≥7 days prior to survey completion were eligible. The incidence and severity of vaccination side effects were assessed.

**Results:** Survey was completed by 2,002 respondents, of whom 26.6% had prior COVID-19 infection (68.8% laboratory confirmed). Prior COVID-19 infection was associated with increased risk of any side effect (risk ratio 1.08, 95% confidence intervals [1.05-1.11]), fever (2.24 [1.86-2.70]), breathlessness (2.05 [1.28-3.29]), flu-like illness (1.78 [1.51-2.10]), fatigue (1.34 [1.20-1.49]) and local reactions (1.10 [1.06-1.15]). It was also associated with increased risk of severe side effects, leading to hospital care (1.56 [1.14-2.12]).

While mRNA vaccines were associated with a higher incidence of any side effect (1.06 [1.01-1.11]) compared to viral vector-based vaccines, these were generally milder (p<0.001), mostly local reactions. Importantly, mRNA vaccine-recipients reported considerably lower incidence of systemic reactions (RR<0.6) including anaphylaxis, swelling, flu-like illness, breathlessness and fatigue, and of side effects requiring hospital care (0.42 [0.31-0.58]).

**Conclusion:** For the first time, our study links prior COVID-19 illness with increased incidence of vaccination side effects and demonstrates that mRNA vaccines cause milder, less frequent systemic side effects, but more local reactions.

**Key messages:**

– People with prior COVID-19 illness appear to experience significantly increased incidence and severity of side effects after receiving the COVID-19 vaccine.
– In this first head-to-head comparison of the safety and reactogenicity of different types of vaccines, it was demonstrated that mRNA vaccines cause milder, less frequent systemic side effects, compared to viral vector vaccines, but more local reactions.

**Tweetable Summary:** A survey of >2000 COVID-19 vaccine-recipients links prior COVID-19 illness with increased incidence of vaccination side effects; mRNA vaccines cause milder, less frequent systemic side effects, but more local reactions.

## Introduction

Coronavirus Disease 2019 (COVID-19) rapidly became a leading cause of death, short and long-term morbidity among people over the age of 45^1,2^, posing an unprecedented burden to healthcare systems, with worldwide economic consequences and prolonged lockdowns^3^. Vaccines currently being rolled out are anticipated to significantly modify these trends. While their effectiveness and safety have been proven in recent studies^4,5,6^, data in specific groups remains lacking. Generally, people with a previous history of COVID-19, in whom vaccination is currently advised^7^, were excluded from the clinical trials^4,5,6^. Whilst it is accepted that prior infection with COVID-19 induces natural immunity potentially lasting for at least six months^8^, yet it is unknown if previous infection may be associated with more vaccination side effects. Moreover, the safety and reactogenicity of the different types of vaccines (mRNA or viral vector-based) have not been compared head-to-head. This anonymized international online survey was conducted to compare the safety profiles of available COVID-19 vaccines and evaluate their side effects in different groups of vaccine recipients.

## Methods

This online survey, developed in plain English language and piloted by experts and lay people, captured basic epidemiological data, details on COVID-19 exposure, vaccination history, the incidence and severity (table e1) of the respective side effects. More specifically, we have enquired about the following symptoms: Localized reactions (pain, swelling, tenderness, redness, itching or other), fever, skin rash, shortness of breath, tingling in the mouth, face, body / extremities, swelling in the face or mouth, generalized swelling, anaphylaxis (severe allergic reaction with face swelling and breathlessness), tiredness or fatigue, flu-like illness, or any other side effects. It was launched via Google Forms on 3^rd^ February 2021, for 11 days, and was shared within the investigators’ institutions, through professional contacts and social media. The only inclusion criterion was the receipt of the first dose of any COVID-19 vaccine at least 7 days prior to survey completion.

The main objectives were to evaluate differences in the incidence and severity of vaccination side effects among (i) people with versus without previously reported COVID-19 infection and (ii) those who received different vaccine types. Moreover, we explored differences in self-reported side effects between the first and second vaccine dose, among different ethnicities and among those with different preconceptions towards the vaccine. Finally, we explored the impact of the interval between COVID-19 exposure and vaccination and the incidence of side effects.

For our main analysis, a positive COVID-19 history was considered in cases of (a) a self-reported history of symptoms consistent with COVID-19 disease, provided that COVID-19 was not excluded by a negative PCR test, (b) a positive COVID-19 PCR test, or (c) a positive COVID-19 antigen test. In a sensitivity analysis, COVID-19 infection was only considered valid if it was confirmed by PCR or antigen testing, while patients with uncertain exposure (clinical history not confirmed by laboratory testing) were excluded.

Between-group differences were assessed using chi-squared and Mann-Whitney U tests for dichotomous and continuous variables, respectively, after Shapiro-Wilk test excluded normal distribution of the latter. Between group differences in the incidence of side effects are presented as risk ratios (RR) with the respective 95% confidence intervals (CI). Predictors of the incidence and severity of side effects were evaluated in univariate, followed by multivariate binomial logistic regression and cumulative link models for ordinal data, respectively. Age, gender, ethnicity, vaccine type, prophylactic analgesia or other medication use prior to vaccination, vaccine preconceptions, and prior COVID-19 exposure were evaluated as potential confounding factors. Unless otherwise specified, the analyses were based on side effect profiles from the first dose of the vaccine.

Ethics approval was not necessary for this anonymized survey.

## Results

Within 11 days, this international online survey was completed by 2,002 participants (table e2, figure e1), mostly health professionals of a working age (median: 45, IQR: 35-50 years). 532 (26.6%) had history of previous COVID-19 infection, of whom 366 (68.8%) were confirmed by PCR (n=273) and/or antigen testing (n=162). COVID-19 infection preceded the first vaccination dose by a median of 87 [IQR: 47-223] days. The majority of respondents were Caucasians (88.3%), mostly from the UK (78.6%) and Greece (16.6%). As anticipated, prior history of COVID-19 infection was more prevalent among frontline workers, health professionals and people from the UK, where a very high incidence of COVID-19 was documented^9^. Moreover, recipients of a viral vector-based vaccine (mainly the AstraZeneca vaccine) were relatively older (figure e2, p<0.001) and were mostly based in the UK (89.7%, compared to 76.4% of those that received viral mRNA vaccines, p<0.001). Finally, doctors were more likely to have received mRNA based vaccine compared to the other groups (p<0.001).

Prior COVID-19 infection was associated with a 8% increase in the risk of having any side effects after the first vaccine dose (RR 1.08, 95% CI [1.05-1.11], table 1, figure 1). We also observed significantly increased risk of self-reported fever (2.24 [1.86-2.70]), breathlessness (2.05 [1.28-3.29]), flu-like illness (1.78 [1.51-2.10]), fatigue (1.34 [1.2-1.49]), local reactions (1.10 [1.06-1.15]) and “other” side effects (1.46 [1.16-1.82]). Among those experiencing side effects, prior COVID-19 infection was associated with increased severity of any side effect, local side effects, or fatigue (p<0.001). More importantly, prior COVID-19 infection was associated with the risk of experiencing a severe side effect, requiring hospital care (1.56 [1.14-2.12]). These observations remained significant in multivariate analyses and our sensitivity analysis (table e3). A similar increase in the risk of any side effects following the second dose in those with prior COVID-19 infection was also noted (1.08 [1.05-1.11]), although the lack of significant associations with specific side effects may result from the limited sample included in this analysis.

**Table 1:** Differences in the incidence and severity of side effects after the first dose of the COVID-19 vaccine among participants who had, or did not have prior COVID-19 infection.

| Side effect | Incidence of side effects: Risk ratio [95% CI] | Incidence of side effects: Multivariate logistic regression, coefficient (p-value) | Severity of side effects: Univariate cumulative risk models (p-value) | Severity of side effects: Multivariate cumulative risk models (p-value) |
| --- | --- | --- | --- | --- |
| Any side effect | 1.08 [1.05-1.11] | 0.575 (0.004) | <0.001 | <0.001 |
| Localized reaction | 1.10 [1.06-1.15] | 0.45 (0.003) | <0.001 | 0.003 |
| Fever | 2.24 [1.86-2.70] | 0.876 (<0.001) | NS | NS |
| Flu-like illness | 1.78 [1.51-2.10] | 0.658 (<0.001) | NS | NS |
| Shortness of breath | 2.05 [1.28-3.29] | 0.651 (0.011) | NS | NS |
| Skin rash | 1.04 [0.54-2.00] | NS | NS | NS |
| Tingling | 1.26 [0.83-1.91] | NS | NS | NS |
| Swelling | 1.00 [0.32-3.14] | NS | NS | NS |
| Generalized Swelling | 1.84 [0.94-3.60] | NS | NS | NS |
| Anaphylaxis | 0.55 [0.06-4.72] | NS | NS | NS |
| Fatigue or Tiredness | 1.34 [1.2-1.49] | 0.418 (<0.001) | <0.001 | <0.001 |
| Other | 1.46 [1.16-1.82] | 0.349 (0.013) | NS | NS |

Worse outcomes associated with prior COVID-19 infection

**Figure 1:**
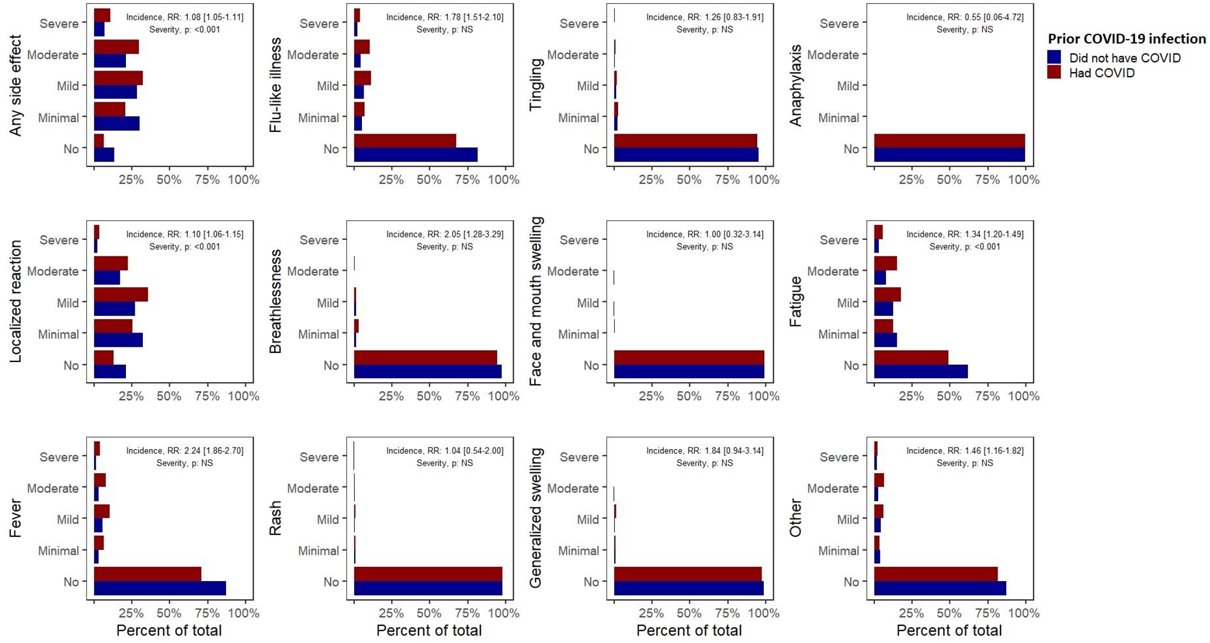
Incidence and severity of self-reported side effects after the first dose of the COVID-19 vaccine among participants who had or did not have known prior COVID-19 infection. Risk ratios less than 1 favours those that did not have prior COVID-19 infection.

Furthermore, significant differences were observed between the side effect profiles of mRNA versus viral vector vaccines (predominantly Pfizer versus AstraZeneca, table 2, figure 2). Overall, recipients of mRNA vaccines reported a higher incidence of any self-reported side effects (1.06 [1.01-1.11]), which however were of significantly milder severity, compared to those who received viral vector vaccines. While mRNA vaccines were associated with an increased incidence of reported local reactions (1.29 [1.19-1.40]), they were associated with considerably lower incidence of self-reported systemic side effects including anaphylaxis (0.19 [0.04-0.62]), fever (0.28 [0.24-0.34]), swelling in the face or mouth (0.29 [0.10-0.80]) or generalized swelling (0.29 [0.15-0.56]), flu-like illness (0.34 [0.29-0.40]), breathlessness (0.43 [0.26-0.70]), fatigue (0.56 [0.51-0.62]) or other side effects (0.67 [0.52-0.86]). These observations were corroborated by multivariate analyses. Most importantly, mRNA vaccines were associated with a significantly lower incidence of severe side effects (requiring hospital care, RR 0.42 [0.31-0.58]).

**Table 2:**
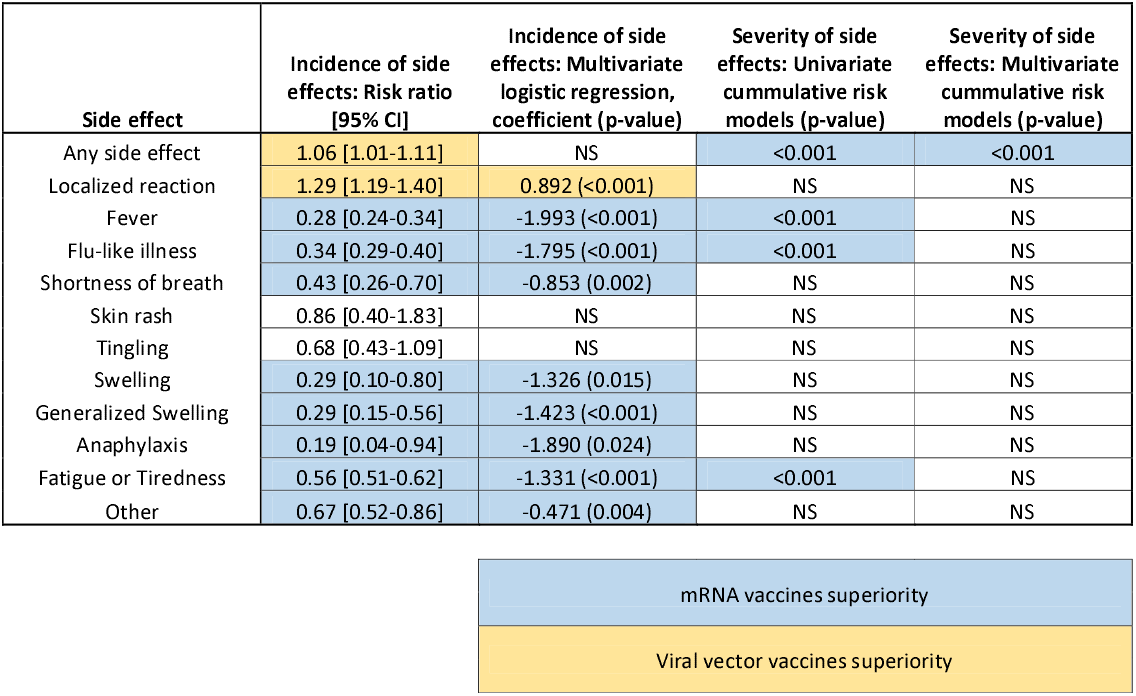
Differences in the incidence and severity of side effects among people who received an mRNA or a viral vector vaccine.

| Side effect | Incidence of side effects: Risk ratio [95% CI] | Incidence of side effects: Multivariate logistic regression, coefficient (p-value) | Severity of side effects: Univariate cumulative risk models (p-value) | Severity of side effects: Multivariate cumulative risk models (p-value) |
| --- | --- | --- | --- | --- |
| Any side effect | 1.06 [1.01-1.11] | NS | <0.001 | <0.001 |
| Localized reaction | 1.29 [1.19-1.40] | 0.892 (<0.001) | NS | NS |
| Fever | 0.28 [0.24-0.34] | -1.993 (<0.001) | <0.001 | NS |
| Flu-like illness | 0.34 [0.29-0.40] | -1.795 (<0.001) | <0.001 | NS |
| Shortness of breath | 0.43 [0.26-0.70] | -0.853 (0.002) | NS | NS |
| Skin rash | 0.86 [0.40-1.83] | NS | NS | NS |
| Tingling | 0.68 [0.43-1.09] | NS | NS | NS |
| Swelling | 0.29 [0.10-0.80] | -1.326 (0.015) | NS | NS |
| Generalized Swelling | 0.29 [0.15-0.56] | -1.423 (<0.001) | NS | NS |
| Anaphylaxis | 0.19 [0.04-0.94] | -1.890 (0.024) | NS | NS |
| Fatigue or Tiredness | 0.56 [0.51-0.62] | -1.331 (<0.001) | <0.001 | NS |
| Other | 0.67 [0.52-0.86] | -0.471 (0.004) | NS | NS |

|  |
| --- |
| mRNA vaccines superiority |
| Viral vector vaccines superiority |

**Figure 2:**
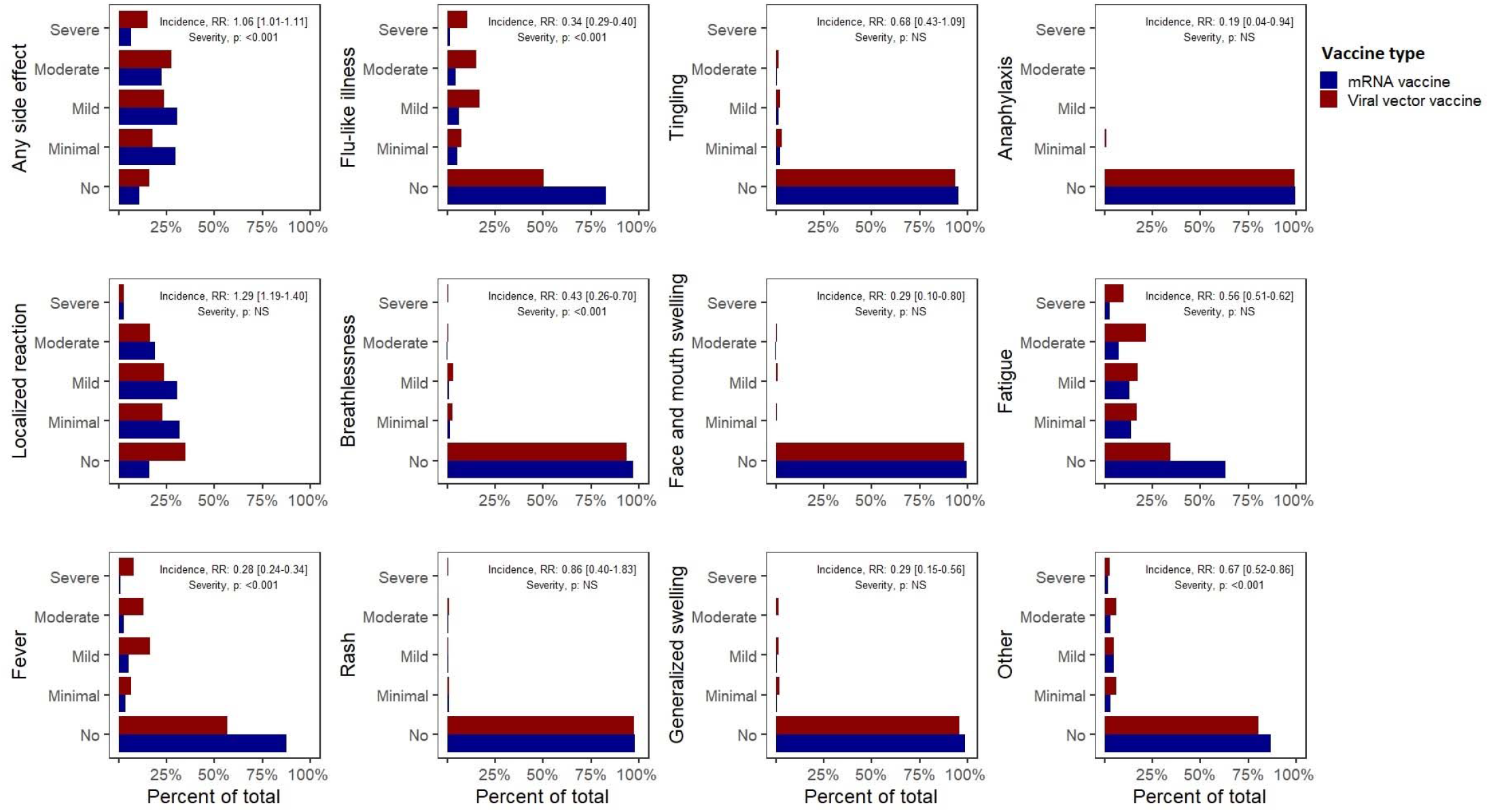
Incidence and severity of side effects after the first dose of an (1) mRNA or (2) viral vector vaccine. Risk ratios less than 1 favours the mRNA vaccines.

In general, the second dose of the vaccine was associated with higher incidence of side effects (table 3). More specifically, respondents reported experiencing more frequently any side effects (1.04 [1.01-1.07]), skin rash (2.25 [1.4-3.62]), fever (1.72 [1.46-2.02]), flu-like illness (1.67 [1.45-1.91]), and fatigue (1.40 [1.28-1.53]). In addition, multivariate regression demonstrated that participants who had side effects after the first vaccine dose, were at significantly higher risk of having the same side effects after the second dose. Among those experiencing side effects, the severity did not significantly differ between the two doses. However, the likelihood of having a severe side effect, requiring hospital care was significantly decreased (0.58 [0.38-0.88]).

**Table 3:** Differences in the incidence and severity of side effects after the second or the first dose of the vaccine.

| Side effect | Incidence of side effects: Risk ratio [95% CI] | Incidence of side effects: Multivariate logistic regression, coefficient (p-value) | Severity of side effects: Univariate cumulative risk models (p-value) | Severity of side effects: Multivariate cumulative risk models (p-value) |
| --- | --- | --- | --- | --- |
| Any side effect | 1.04 [1.01-1.07] | NS | NS | NS |
| Localized reaction | 0.98 [0.94-1.03] | 2.469 (<0.001) | NS | NS |
| Fever | 1.72 [1.46-2.02] | 1.3 (<0.001) | NS | NS |
| Flu-like illness | 1.67 [1.45-1.91] | 0.979 (0.001) | NS | NS |
| Shortness of breath | 0.95 [0.57-1.61] | 4.491 (<0.001) | NS | NS |
| Skin rash | 2.25 [1.4-3.62] | 4.297 (<0.001) | 0.05 | NS |
| Tingling | 1.31 [0.89-1.92] | 3.096 (<0.001) | NS | NS |
| Swelling | 2.03 [0.87-4.77] | NS | NS | NS |
| Generalized Swelling | 1.2 [0.61-2.34] | 4.925 (<0.001) | NS | NS |
| Anaphylaxis | 2.54 [0.72-8.98] | 4.747 (0.012) | NS | NS |
| Fatigue or Tiredness | 1.4 [1.28-1.53] | 0.868 (<0.001) | NS | NS |
| Other | 1.05 [0.83-1.32] | 2.104 (<0.001) | NS | NS |

Worse outcomes after the second COVID-19 vaccine dose

Stratification by ethnicity revealed that white participants reported lower incidence of fever (0.62 [0.48-0.79]) and flu-like illness (0.78 [0.62-0.97]), compared to the remaining participants (table e4). Finally, those reporting pre-vaccination concern about the safety of the vaccine, reported more often tingling (2.23 [1.45-3.42]), breathlessness (1.73 [1.00-2.98]), and fatigue (1.17 [1.03-1.34], table e5).

Multivariate analyses also revealed a strong, negative association between age and the self-reporting of any side effect, local reactions, fever, flu-like illness, rash, tingling, generalized swelling and fatigue (p<0.01). Finally, a history of allergy was associated with an increased incidence of self-reported breathlessness and rash (p<0.01). However, as described in the previous paragraphs and tables, most of the associations observed in univariate analyses remained significant in multivariate analyses accounting for these and other potential confounding factors.

## Discussion

People with prior COVID-19 exposure were largely excluded from the vaccine trials^4,5,6^ and, as a result, the safety and reactogenicity of the vaccines in this population have not been previously fully evaluated. For the first time, this study demonstrates a significant association between prior COVID-19 infection and a significantly higher incidence and severity of self-reported side effects after vaccination for COVID-19. Consistently, compared to the first dose of the vaccine, we found an increased incidence and severity of self-reported side effects after the second dose, when recipients had been previously exposed to viral antigen. In view of the rapidly accumulating data demonstrating that COVID-19 survivors generally have adequate natural immunity for at least 6 months, it may be appropriate to re-evaluate the recommendation for immediate vaccination of this group.

Moreover, this is the first head-to-head real-world comparison of the self-reported safety of viral vector versus mRNA vaccines, with the latter associated with a 58% decreased incidence of self-reported severe side effects, requiring hospital care. While more recipients of mRNA vaccines reported at least one (any) side effect, the difference was predominantly driven by the frequent local reactions, while the incidence of each of the systemic side effects evaluated, which are more burdensome to the recipients, was significantly reduced. Recipients of the viral vector-based vaccines were relatively older. However, differences in the incidence of adverse events were confirmed in multivariate analyses accounting for the age of the respondents as a covariate. Moreover, given that older people reported side effects less frequently, potential bias due to age difference would be expected to favour viral vector-based vaccines. These findings may have an impact on vaccine choice, and health policies.

The main strengths of our study include a large study population that better reflects real-life compared to the populations studied in the clinical trials, the availability of adequate details about the participants and the vaccines’ safety profiles, and very limited missing data. Potential respondents bias is the main limitation of any survey and since this survey was shared though social media, we were not able to estimate the non-response rate. However, respondents bias is more likely to affect the absolute incidence of side effects, that we did not evaluate here, rather than the relative incidence and severity across different groups of people. Potential recall bias should also be mentioned, although all participants had been vaccinated within 10 weeks prior to completing the survey. As noted, most respondents were from the UK and Greece due to the ability of the investigators to establish contacts quickly to publicise this survey. The UK has also been successful in rolling out COVID-19 vaccines quickly leading to more of those invited being eligible to participate. It is not surprising that Pfizer vaccine was the most delivered vaccine as it was the first vaccine to be licensed within the UK, with more individuals receiving it in total when the survey was circulated.

In conclusion, this extensive survey of over 2,000 recipients of the COVID-19 vaccines links previous COVID-19 illness with increased incidence of vaccination side effects. It also demonstrates that mRNA vaccines cause milder, less frequent systemic side effects, but more local reactions. These findings will need to be validated in clinical studies, preferably randomized controlled trials, including patients from multiple groups.

## Data Availability

Access to respondents level data is available upon request to the authors.

## Acknowledgements

AGM was supported by the NIHR Manchester Biomedical Research Centre (NIHR Manchester BRC). We are thankful to Dr. Matthew Snape (Oxford Vaccine Group, University of Oxford) for his advice regarding vaccine safety and reactogenicity tracking, and to the Coronavirus Medical Group Greece for disseminating our survey among Greek health professionals.

## ONLINE APPENDIX

**Table e1:**
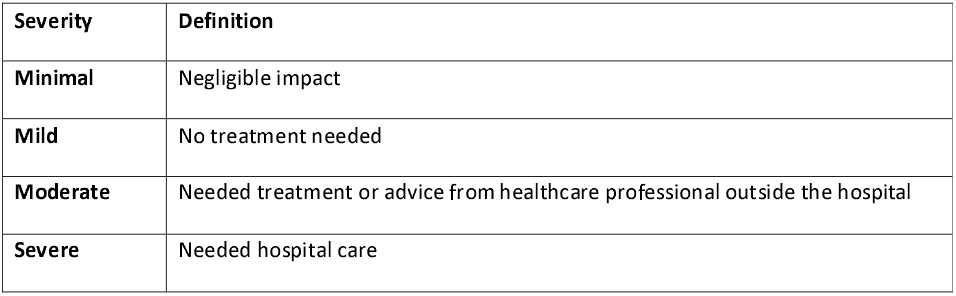
Definitions of side effects severity.

**Table e2:** Baseline characteristics of study participants. Continuous variables as presented as medians [IQR] and categorical as n (%). Between group differences were anticipated and explained by the incidence of COVID-19 in different subgroups. Characteristically, higher incidence of prior COVID-19 infection was observed among frontline workers, health professionals and among British people (a very high incidence of COVID-19 was documented in the UK).

| **Characteristics** | **Participants with prior COVID-19 infection (n=532)** | **Participants with no known prior COVID-19 infection (n=1,470)** | **Missing data** | **Between group differences (P-value)** |
| --- | --- | --- | --- | --- |
| **Gender (Female)** | 393 | 1,051 | 0.7% | NS |
| **Weight (kg)** | 75 [64-88] | 74 [64-85] | 4.0% | NS |
| **Height (cm)** | 168 [163-173] | 168 [162-175] | 2.2% | NS |
| **Country** |  |  | 0.6% | <0.001 |
| Europe |  |  |  |  |
| UK | 472 (88.7%) | 1,100 (74.8%) |  |  |
| Greece | 38 (7.1%) | 294 (20%) |  |  |
| Other European countries | 10 (1.9%) | 30 (2.0%) |  |  |
| Americas | 5 (0.9%) | 17 (1.2%) |  |  |
| Asia | 5 (0.9%) | 17 (1.2%) |  |  |
| Africa | 0 (0%) | 1 (0.1%) |  |  |
| **Ethnicity** |  |  | 1.8% | NS |
| White | 464 (87.2%) | 1,303 (88.6%) |  |  |
| Asian | 35 (6.6%) | 63 (4.3%) |  |  |
| Arab | 21 (3.9%) | 45 (3.1%) |  |  |
| Other | 7 (1.3%) | 28 (1.9%) |  |  |
| **Role** |  |  | 3.2% | <0.001 |
| Doctor | 140 (26.3%) | 486 (33.1%) |  |  |
| Nurse | 125 (23.5%) | 188 (12.8%) |  |  |
| Other health professional | 161 (30.3%) | 382 (26.0%) |  |  |
| Not a health professional | 105 (19.7%) | 401 (27.8%) |  |  |
| **Frontline workers** | 372 (69.9%) | 795 (54.1%) | 0.6% | <0.001 |
| **COVID-19 prior to vaccination** |  |  | 0% |  |
| Laboratory confirmed exposure | 366 (68.8%) | NA |  |  |
| Consistent symptoms, not tested | 166 (31.2%) | NA |  |  |
| No known exposure | NA | 1,470 (100%) |  |  |
| **Vaccine type** |  |  | 0.5% | NS |
| Pfizer | 443 (83.3%) | 1,230 (83.7%) |  |  |
| Oxford – AstraZeneca | 80 (15.0%) | 202 (13.7%) |  |  |
| Other | 4 (0.8%) | 20 (1.4%) |  |  |
| Unknown | 2 (0.4%) | 3 (0.2%) |  |  |
| **Vaccine preconception** |  |  | 0.8% | NS |
| Positive | 343 (64.5%) | 1,027 (69.9%) |  |  |
| Neutral | 76 (14.3%) | 174 (11.8%) |  |  |
| Negative | 110 (20.7%) | 259 (17.6%) |  |  |
| **Second vaccine dose received** | 114 (21.4%) | 411 (28.0%) | 0% | 0.004 |
| **Past Medical History** |  |  | 7.7% |  |
| Chronic Cardiac Disease | 9 (1.7%) | 25 (1.7%) |  | NS |
| Chronic Respiratory Disease | 74 (13.9%) | 171 (11.6%) |  | NS |
| Chronic Kidney Disease | 4 (0.8%) | 9 (0.6%) |  | NS |
| Chronic Liver Disease | 1 (0.2%) | 6 (0.4%) |  | NS |
| Chronic Neurological Disease | 8 (1.5%) | 17 (1.2%) |  | NS |
| Active cancer | 1 (0.2%) | 9 (0.6%) |  | NS |
| Asplenia | 1 (0.2%) | 4 (0.3%) |  | NS |
| Allergy | 56 (10.5%) | 134 (9.1%) |  | NS |
| Diabetes | 17 (3.2%) | 49 (3.3%) |  | NS |
| Hay fever, eczema | 114 (21.4%) | 251 (17.1%) |  | 0.04 |
| Immunosuppression | 14 (2.6%) | 49 (33.3%) |  | NS |
| Transplantation history | 0 (0%) | 0 (0%) |  | NS |
| None | 282 (53.0%) | 825 (56.1%) |  | NS |

*\* Participants with prior COVID-19 exposure were younger compared to those without prior exposure.*

**Table e3:**
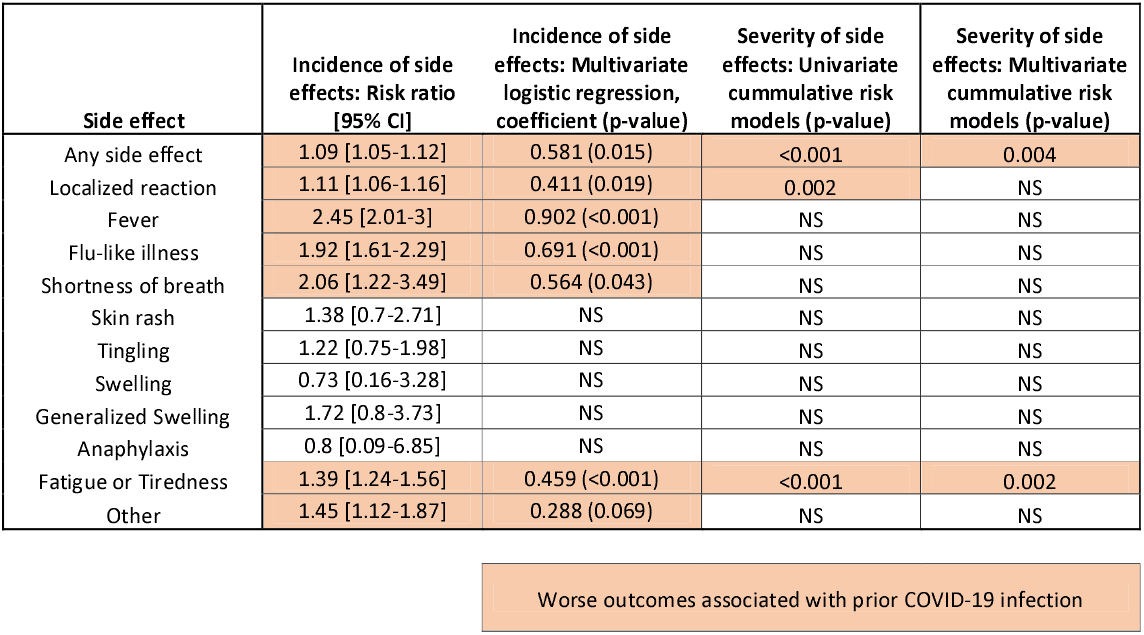
Differences in the incidence and severity of side effects after the first dose of the COVID-19 vaccine among participants who had or did not have prior self-reported COVID-19 infection. Sensitivity analysis only including participants with prior COVID-19 infection confirmed with a consistent PCR or antibody test (n=366) versus those without any suspicion of prior COVID-19 infection (n=1,470).

**Table e4:**
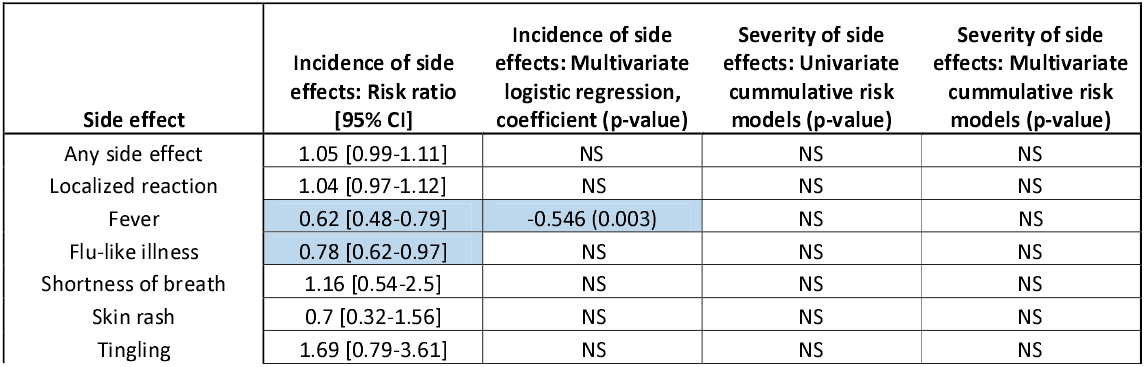
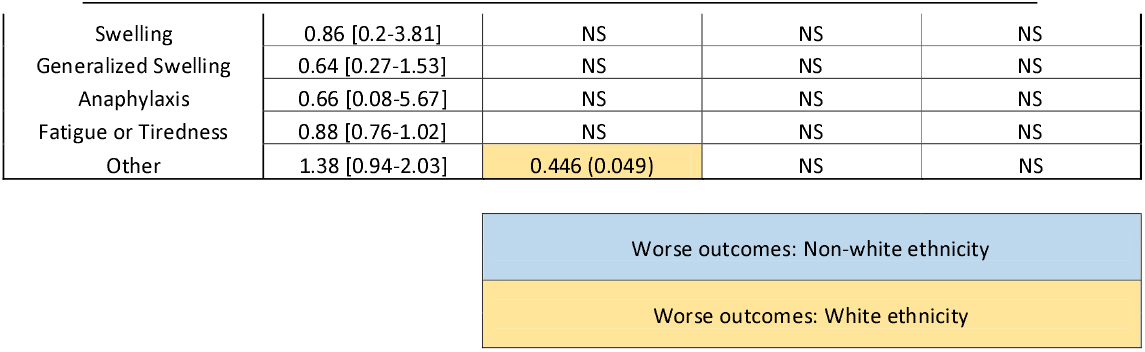
Differences in the incidence and severity of side effects among different ethnicities (white or other).

Worse outcomes associated with prior COVID-19 infection **Table e4:** Differences in the incidence and severity of side effects among different ethnicities (white or other).
| Side effect | Incidence of side effects: Risk ratio [95% CI] | Incidence of side effects: Multivariate logistic regression, coefficient (p-value) | Severity of side effects: Univariate cumulative risk models (p-value) | Severity of side effects: Multivariate cumulative risk models (p-value) |
| --- | --- | --- | --- | --- |
| Any side effect | 1.05 [0.99-1.11] | NS | NS | NS |
| Localized reaction | 1.04 [0.97-1.12] | NS | NS | NS |
| Fever | 0.62 [0.48-0.79] | -0.546 (0.003) | NS | NS |
| Flu-like illness | 0.78 [0.62-0.97] | NS | NS | NS |
| Shortness of breath | 1.16 [0.54-2.5] | NS | NS | NS |
| Skin rash | 0.7 [0.32-1.56] | NS | NS | NS |
| Tingling | 1.69 [0.79-3.61] | NS | NS | NS |
| Swelling | 0.86 [0.2-3.81] | NS | NS | NS |
| Generalized Swelling | 0.64 [0.27-1.53] | NS | NS | NS |
| Anaphylaxis | 0.66 [0.08-5.67] | NS | NS | NS |
| Fatigue or Tiredness | 0.88 [0.76-1.02] | NS | NS | NS |
| Other | 1.38 [0.94-2.03] | 0.446 (0.049) | NS | NS |

|  |
| --- |
| Worse outcomes: Non-white ethnicity |
| Worse outcomes: White ethnicity |

**Table e5:**
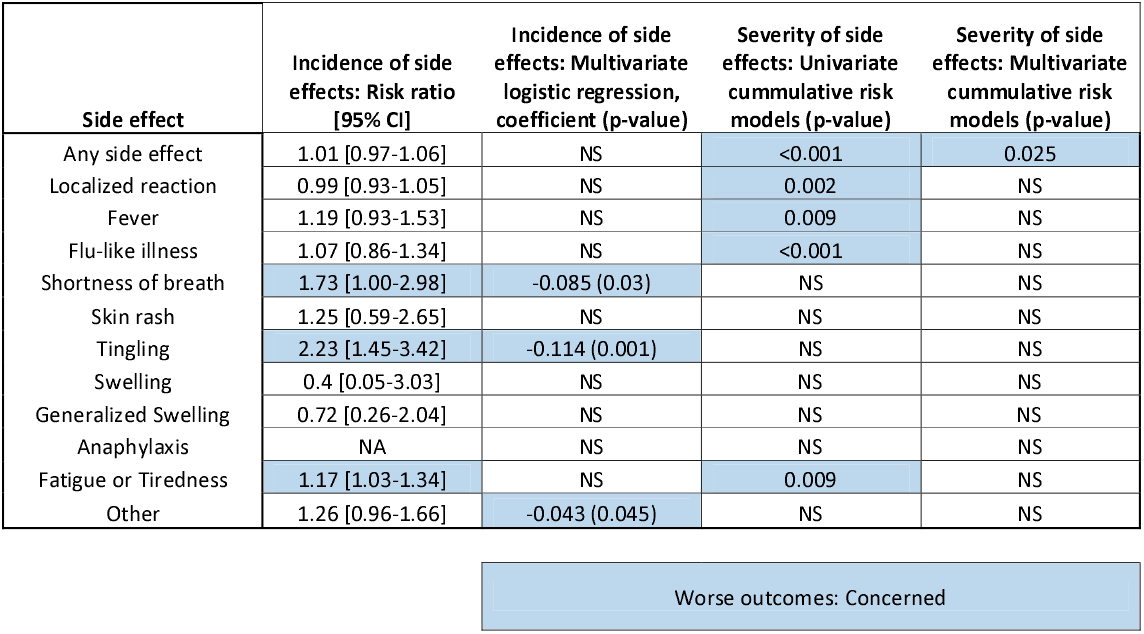
Differences in the incidence and severity of side effects among people with different preconception toward the vaccine prior to vaccination, those who were keen to receive the vaccine versus those who were concerned about receiving the vaccine.

| Side effect | Incidence of side effects: Risk ratio [95% CI] | Incidence of side effects: Multivariate logistic regression, coefficient (p-value) | Severity of side effects: Univariate cumulative risk models (p-value) | Severity of side effects: Multivariate cumulative risk models (p-value) |
| --- | --- | --- | --- | --- |
| Any side effect | 1.01 [0.97-1.06] | NS | <0.001 | 0.025 |
| Localized reaction | 0.99 [0.93-1.05] | NS | 0.002 | NS |
| Fever | 1.19 [0.93-1.53] | NS | 0.009 | NS |
| Flu-like illness | 1.07 [0.86-1.34] | NS | <0.001 | NS |
| Shortness of breath | 1.73 [1.00-2.98] | -0.085 (0.03) | NS | NS |
| Skin rash | 1.25 [0.59-2.65] | NS | NS | NS |
| Tingling | 2.23 [1.45-3.42] | -0.114 (0.001) | NS | NS |
| Swelling | 0.4 [0.05-3.03] | NS | NS | NS |
| Generalized Swelling | 0.72 [0.26-2.04] | NS | NS | NS |
| Anaphylaxis | NA | NS | NS | NS |
| Fatigue or Tiredness | 1.17 [1.03-1.34] | NS | 0.009 | NS |
| Other | 1.26 [0.96-1.66] | -0.043 (0.045) | NS | NS |

|  |
| --- |
| Worse outcomes: Concerned |

**Figure e1:**
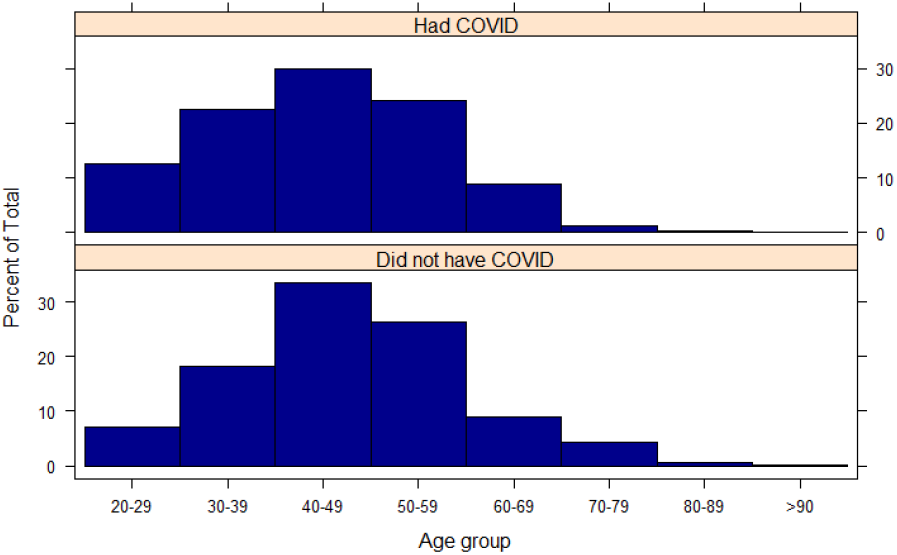
Age of the participants, stratified by whether they had or did not have a previous COVID-19 infection.

**Figure e2:**
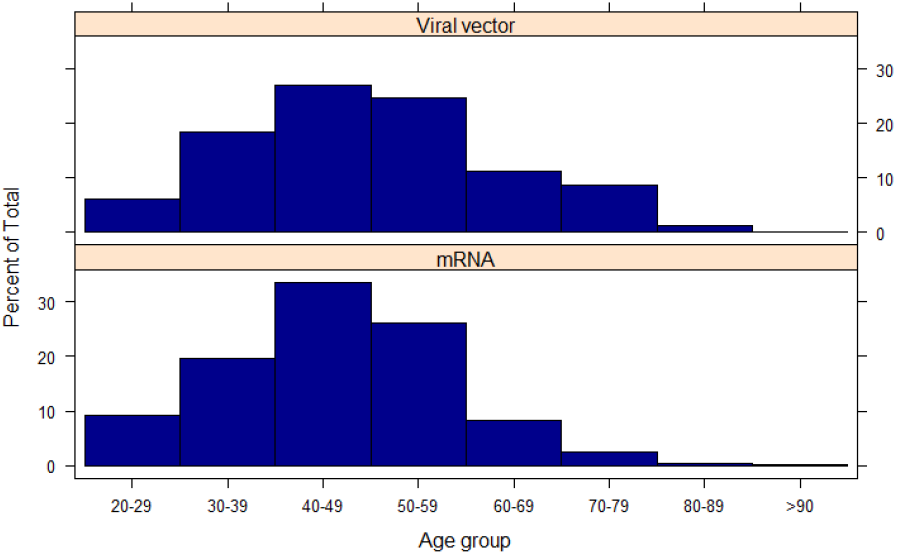
Age of the participants, stratified by the type of vaccine they received

